# Initial Model for USA CoVID-19 Resurgence

**DOI:** 10.1101/2020.09.16.20196063

**Authors:** Genghmun Eng

## Abstract

Early CoVID-19 growth obeys: 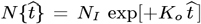, with *K*_*o*_ = [(ln 2)/(*t*_*dbl*_)], where t_*dbl*_ is the pandemic growth *doubling time*. Given 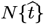, the daily number of new CoVID-19 cases is 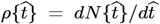. Implementing society-wide *Social Distancing* increases the *t*_*dbl*_ *doubling time*, and a linear function of time for *t*_*dbl*_ was used in our *Initial Model*:

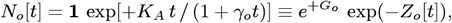

to describe these changes, where the [t]-axis is time-shifted from the 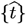–axis back to the pandemic start, and *G*_*o*_ ≡ [*K*_*A*_/ *γ*_*o*_]. While this N_*o*_[t] successfully modeled the USA CoVID-19 progress from 3/2020 to 6/2020, this equation could not easily model some quickly decreasing *ρ*[*t*] cases (“*fast pandemic shutoff* “), indicating that a second process was involved. This situation was most evident in the initial CoVID-19 data from *China, South Korea*, and *Italy*. Modifying *Z*_*o*_[*t*] to allow exponential cutoffs:

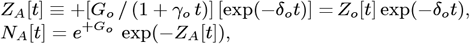

resulted in an *Enhanced Initial Model (EIM)* that significantly improved data fits for these cases.

After 6/2020, many regions of the USA “*opened up*”, loosening their *Social Distancing* requirements, which led to a sudden USA CoVID-19 *Resurgence*. Extrapolating the USA *N*_*o*_[*t*] 3/2020-6/2020 results to 9/2020 as an *Initial Model Baseline (IMB)*, and subtracting this *IMB* from the newer USA data gives a *Resurgence Only* function, which is analyzed here. This USA CoVID-19 *Resurgence* function differs significantly from the N_*o*_[t] IMB functional form, but it was well-modeled by the *N*_*A*_[*t*] *fast pandemic shutoff* function. These results indicate that: (a) the gradual increase in *t*_*dbl*_ *doubling time* from society-wide shut-downs is likely due to eliminating of a large number of population gathering points that could have enabled CoVID-19 spread; and (b) having a non-zero *δ* _*o*_ *fast pandemic shutoff* is likely due to more people wearing masks more often [with *12 Figures*].

## 1 Introduction

The CoVID-19 pandemic started late in 2019, becoming world-wide in early 2020, with CoVID-19 spread evolving differently in various areas. Many publicly available databases were set up to track the disease, to assist epidemiologists, scientists, and policy makers in visualizing CoVID-19 spread. The widely available *bing*.*com*^**1**^ CoVID-19 database was used here. These databases underpin model projections, allowing quick evaluation of how different inputs affect the predicted outcome. Our goal was to empirically model a wide range of data with a small number of parameters, where different values for these parameters could span the range of observed CoVID-19 evolution among regions.

The 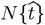 number of CoVID-19 cases starts with an exponential growth:

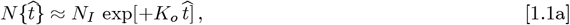

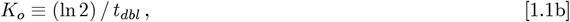

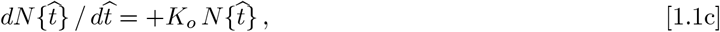

where *N*_*I*_ at time 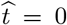 is the number of infected people, *K*_*o*_ is a *rate constant* for how fast an infected person spreads CoVID-19 to others, and *t*_*dbl*_ is the pandemic *doubling time*. This is the basis for a large number of **SEIR** (*Susceptible, Exposed, Infected, and Recovered or Removed*) pandemic models, which are often implemented as systems of local differential equations.

Implementing society-wide measures for non-infected people is inherently a non-local process. How it impacts pandemic spread is often not the main focus of **SEIR** models, which are local. However, when governments mandated *Social Distancing*, starting with shut-down of large-scale gathering places at some 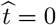 point, the 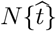 response was fairly quick. Within days, the *t*_*dbl*_ was empirically observed to gradually lengthen, likely due to these shut-downs preventing a large number of people from gathering together and spreading CoVID-19.

Our prior work^**2-3**^ showed that an *Initial Model*^**2**^, with a linear function of time for gradual *t*_*dbl*_ changes:

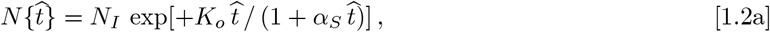

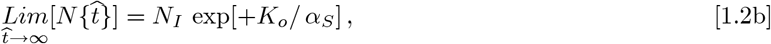

successfully fit a lot of early CoVID-19 pandemic data. More importantly, Eq. [1.2b] showed that this *Initial Model* allows for CoVID-19 pandemic shut-off, prior to infecting the whole population.

An exception was CoVID-19 spread in Italy, having a much faster pandemic shut-off than Eq. [1.2a] predicted. We attributed this to a second CoVID-19 mitigation process that was modeled with a *δ*_*o*_ exponential decay time constant:

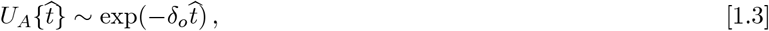

where *δ*_*o*_ = 0 is the absence of this second process. Including this second process^**4**^ gave an *Enhanced Initial Model (EIM)*, which then successfully modeled CoVID-19 spread in Italy.

While the *Initial Model*^**2-3**^ successfully predicted the USA CoVID-19 evolution from March 2020 through early June 2020, widespread “*opening-up*” of various gathering places (such as *local bars* and *hair and nail salons*) in mid-June 2020 created a large-scale USA CoVID-19 *Resurgence*.

A new model for USA CoVID-19 *Resurgence* is developed here. Our prior (3/2020-6/2020) USA CoVID-19 function was used as an *Initial Model Baseline (IMB)*. This *IMB* was projected out to 9/2020, and subtracted from all follow-on USA data, to give a *Resurgence Only* function. As detailed next, the number of USA CoVID-19 *Resurgence* cases can substantially exceed the expected pre-Resurgence total. More importantly, this CoVID-19 *Resurgence* was also found to require a *δ*_*o*_ ≠ 0 *EIM* in order to achieve a good data fit.

This *δ*_*o*_ ≠ 0 result is similar to the prior analysis of CoVID-19 spread in Italy^**4**^. The fact that the *δ*_*o*_ ≠ 0 *EIM* function is needed to model *Resurgence*, instead of an Eq. [1.2a] *IMB* -type function, helps to identify the CoVID-19 second process. After the *Social Distancing* period of 3/20-6/20, new post-6/20 society-wide recommendations or mandates to wear masks were put in place, which likely gives rise to this faster *Resurgence* pandemic shut-off.

## 2 Background

Let 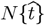 model the total number 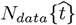 of CoVID-19 cases in a locality, with 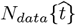 having end-points {*N*_*I*_, *N*_*F*_}. Then 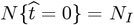, and:

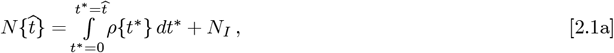

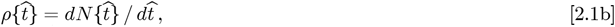

where 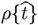 is the predicted number of daily new CoVID-19 cases. Early CoVID-19 growth obeys 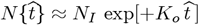, with *K*_*o*_ = [(ln 2)/*t*_*dbl*_], where *t*_*dbl*_ is the pandemic *doubling time*, but if society-wide *Social Distancing* starts at 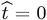, then *t*_*dbl*_ can lengthen for 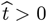. The prior 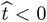 exponential growth phase, before *Social Distancing* started, is not applicable for estimating *Social Distancing* parameters.

Our *Initial Model* for CoVID-19 spread and *t*_*dbl*_ lengthening^**2-3**^ is given in the above Eqs. [1.2a]-[1.2b]. In Eq. [1.2a], different *N*_*I*_ values alter the 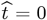 points. However, all these time axes can be shifted to a new *t* = 0 point that estimates the CoVID-19 pandemic start:

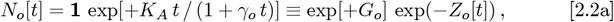

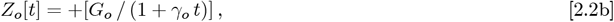

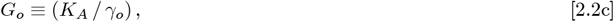

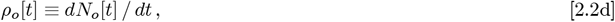

along with the boundary conditions that *N*_*o*_[*t* = *t* _*I*_] *N*_*I*_ and *N*_*o*_[*t* = *t*_*F*_] ≈ *N*_*F*_ occur over the (*t*_*F*_ − *t*_*I*_) time interval. The *t*_*I*_ value is set by the prior {*K*_*o*_, *α*_*s*_} values. Specifically, at 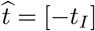, Eq. [1.2b] must give:

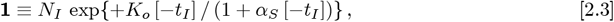

which then individually sets {*t*_*I*_, *t*_*F*_} as follows:

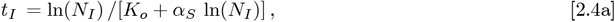

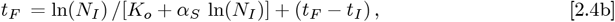

with the {*N*_*I*_, *t* _*I*_, *N*_*F*_, *t* _*F*_} group uniquely determining 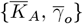:

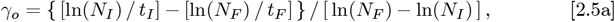

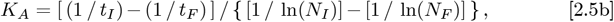

which sets the Eq. [2.2b] *Z*_*o*_[*t*] function. The total number of cases 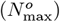 at the pandemic end, and the long-time tail for *α*_*o*_[*t*] are each given by:

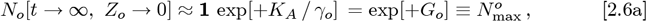

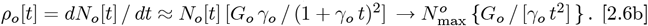

The {0 < *t* < *t*_*I*_} period, prior to *Social Distancing* start, estimates what the pandemic would have looked like, had *Social Distancing* begun at *t* = 0.

Using USA CoVID-19 data from *bing*.*com*^**1**^ from 3/21/2020 through 6/7/2020, we derived the following *Initial Model Baseline (IMB)* best fit as shown in **Figs. 1-2**, using these parameter values:

**Figure 1.**
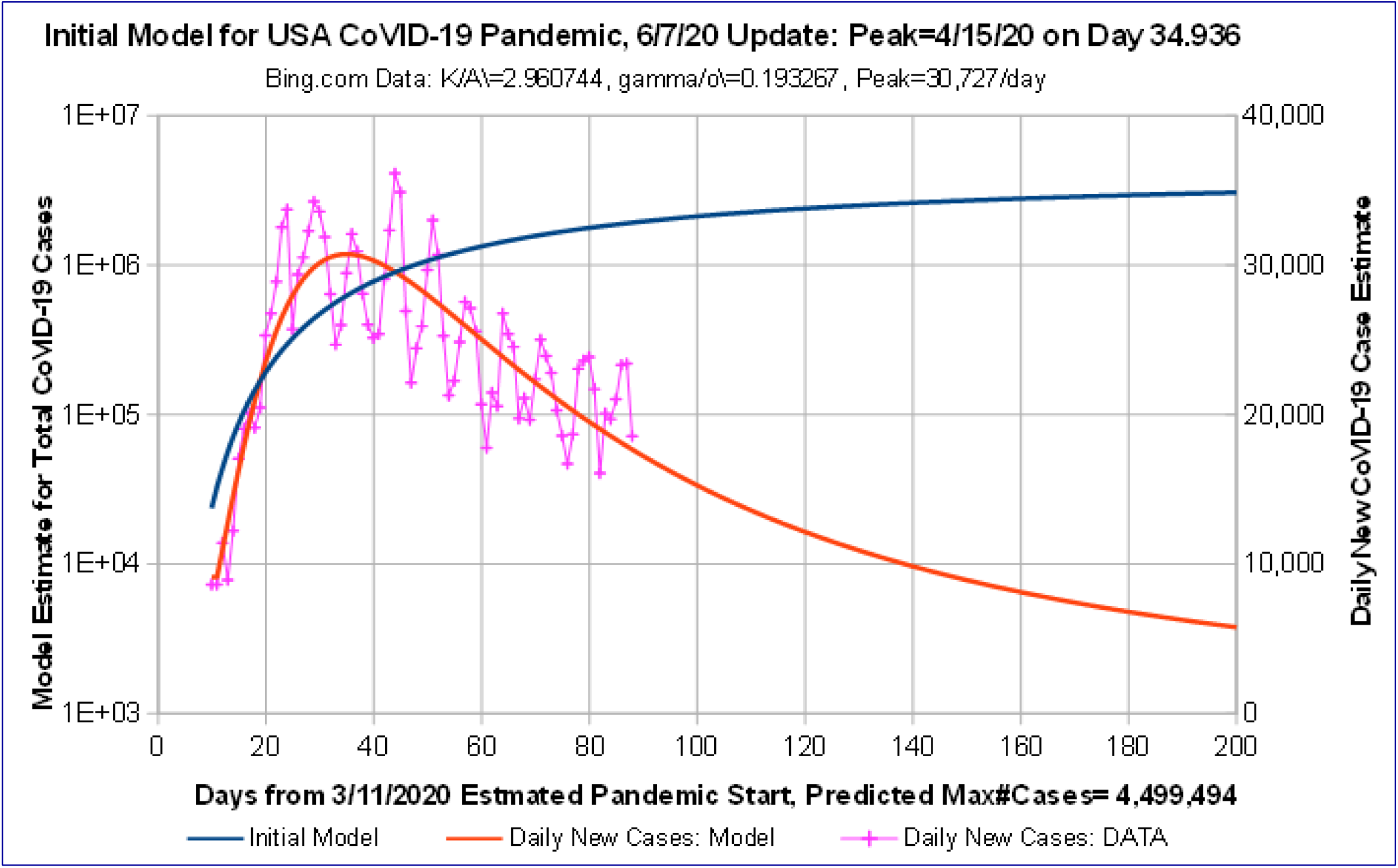
*Initial Model Baseline (IMB):* USA CoVID-19 Projections vs data to 6/7/2020. Predicted Number of Daily CoVID-19 Cases has a peak of 30,727 cases/day on 4/15; with 4,499,494 cases total; and ∼5,783 new cases/day at Day 200 on 9/27/2020.

**Figure 2.**
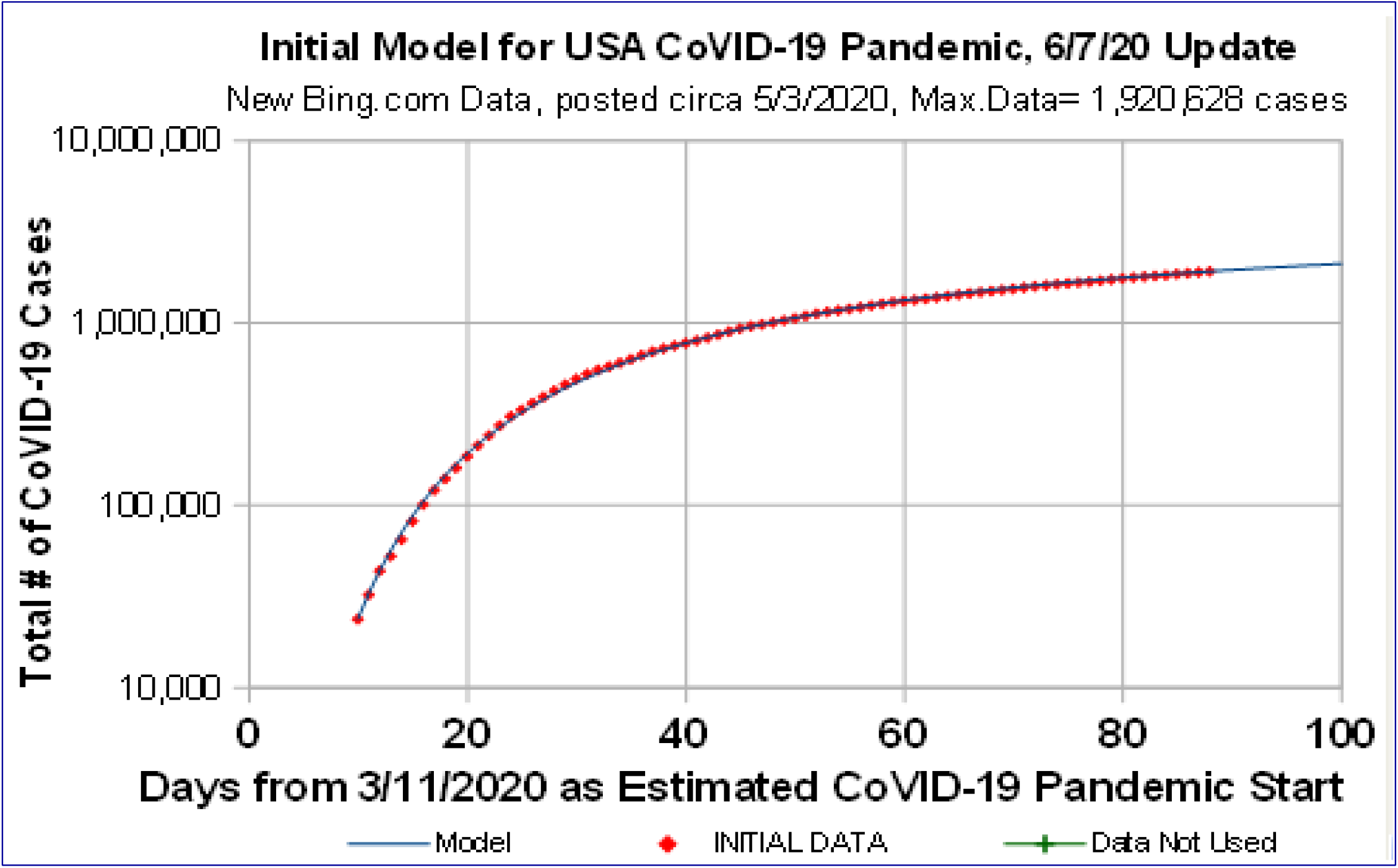
*Initial Model Baseline (IMB)*: USA CoVID-19 Projections vs data to 6/7/2020. Revised *bing*.*com* data, circa 5/3/2020, changed all values back to the pandemic start. *Initial Model* appears to be a good data fit.

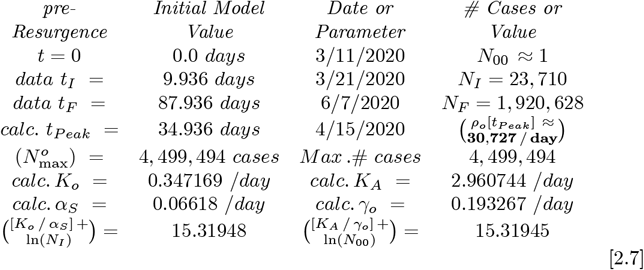

Shortly after 6/7/2020, many states and cities around the USA “*opened up*” nearly simultaneously, loosening *Social Distancing* restrictions. This optimistic action led to a sudden USA CoVID-19 *Resurgence*.

## 3 *Initial Model* for CoVID-19 *Resurgence*

To model CoVID-19 *Resurgence*, the **Figs. 1-2** *IMB* curve values were subtracted from the new USA data totals. When the total number of CoVID-19 *Resurgence cases*, 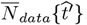, showed a trend above the *IMB* baseline, then 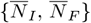 could be used as the 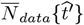 data end-points. Let 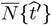 model this CoVID-19 *Resurgence data*, so that 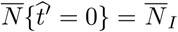. Then:

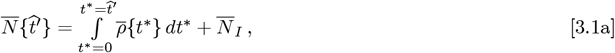

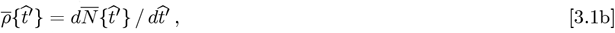

where 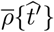 is the predicted number of daily new CoVID-19 *Resurgence* cases. Early CoVID-19 *Resurgence* can obey 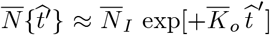, where 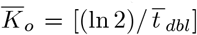 and 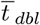 is the pandemic *Resurgence doubling time*. Using an *Initial Resurgence Model (IRM)* that parallels the prior section *IMB* gives:

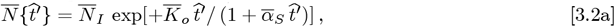

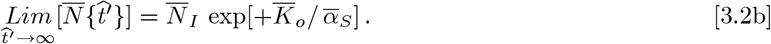

The best 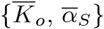 are set by minimizing the *rms* error between the Eq. [3.2a] function and the measured CoVID-19 *Resurgence* data. In Eq. [3.2a], the 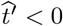 data where 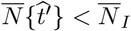, are not used to estimate the 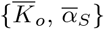 pandemic *Resurgence* parameters. As Eq. [3.2b] shows, this *IRM* allows pandemic shut-off before the CoVID-19 *Resurgence* infects the whole population. In Eq. [3.2a], different 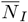 values alter the 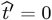 points, but all these time axes can be shifted to a new *t*′ = 0 point that estimates the CoVID-19 *Resurgence* start:

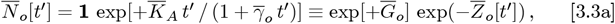

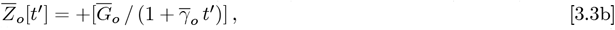

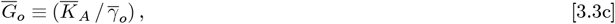

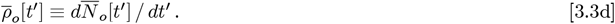

Since the 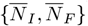 data end points span a time interval of 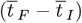, the constraints 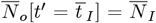 and 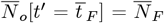 determines 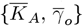 as follows. Using a method similar to Eq. [2.3] and Eqs. [2.4a]-[2.4b], the best fit 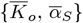 values from Eq. [3.2a] set 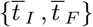:

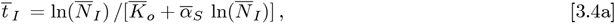

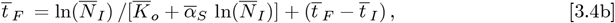

with the 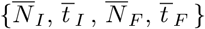 group uniquely determining 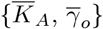:

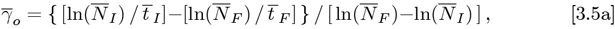

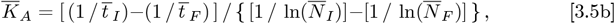

which sets the Eq. [3.3b] 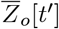 function. The total number of cases 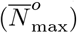 at pandemic end, and the long-term tail for 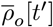 are each given by:

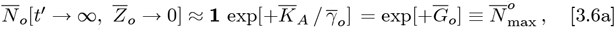

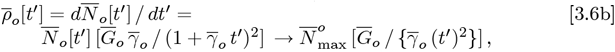

showing that 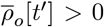. Either 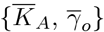 or 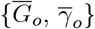 can be used as the primary variables. The *IRM* analysis results for the USA CoVID-19 *Resurgence* are shown in **Figs. 3-4**, using the best-fit 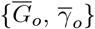 values. The prior 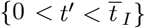 period in **Fig. 3** estimates what the pandemic would have looked like, if *Resurgence Social Distancing* had begun at *t*′ = 0. The best fit parameter values for **Figs. 3-4** are:

**Figure 3.**
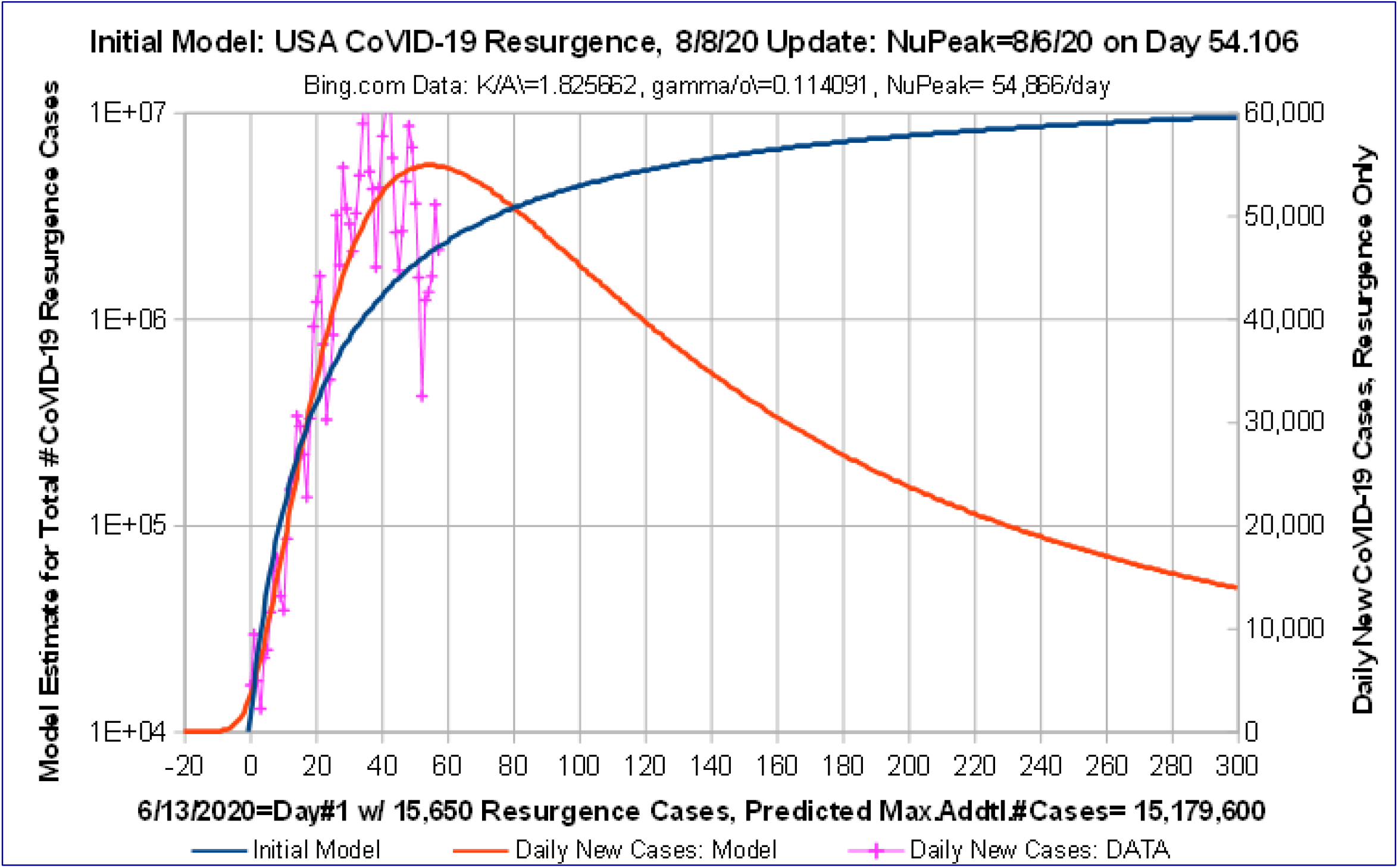
*Initial Model* fit for CoVID-19 *Resurgence:* USA Data 6/13/20 to 8/8/20. The **Fig. 1** *IMB* has data through 6/7/2020. It was extrapolated to 8/27/2020, then subtracted from The actual data to create *Resurgence Only* data, which was then fitted using the *Initial Model*.

**Figure 4.**
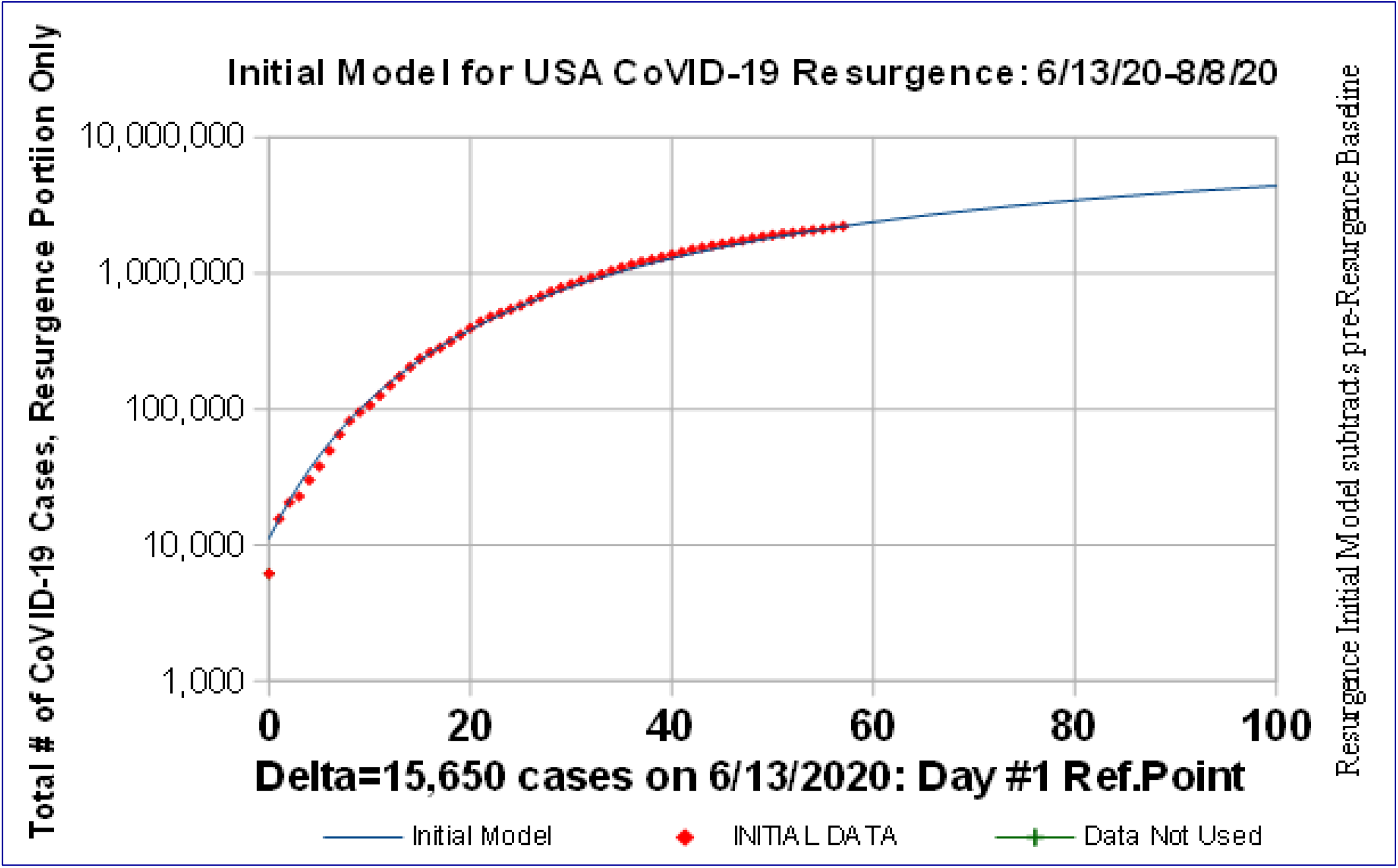
USA CoVID-19 *Resurgence* Only: Data vs *Initial Model* Fit, 6/13/20 to 8/8/20. *Initial Model Baseline (IMB)* was subtracted from actual data to set Resurgence Only Data. Resurgence Start Day #1 was set to 6/13/2020 with ***N=15***,***650*** cases above *IMB*.

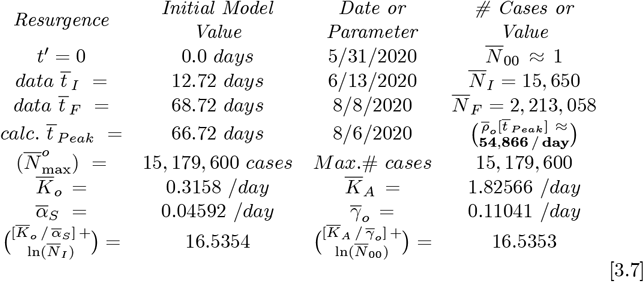

Unfortunately, **Fig. 3** shows that the 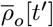 *IRM* data fit is not that good. The 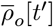 function has a long tail, which overestimates 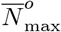, as the maximum number of CoVID-19 *Resurgence* cases at the pandemic end.

## 4 Enhanced Model: USA CoVID-19 *Resurgence*

Since the observed 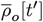 *Resurgence* data decreases much quicker than the *IRM* prediction, the USA CoVID-19 *Resurgence* has a fast pandemic shutoff, which is similar to our prior study^**3-4**^ of Italy CoVID-19 data. That Italy data was most successfully modeled by introducing a second process having an exponential decay in time. Generalizing the *IRM* model of Eq. [3.3a] similarly gives this *Enhanced Initial Model (EIM)* for *Resurgence*, where 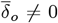 in Eq. [4.1c] characterizes this second process:

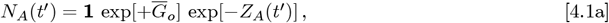

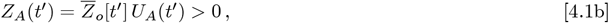

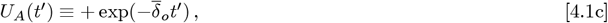

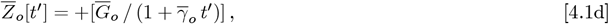

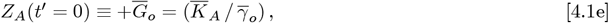

The above Eqs. [4.1a]-[4.1e] have these limits:

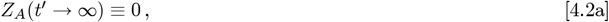

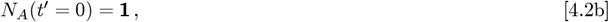

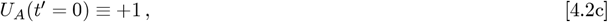

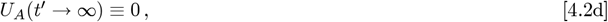

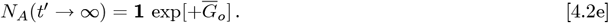

For easier data fitting when *δ*_*o*_ ≠ 0, the Eq. [4.2b] condition that *N*_*A*_(*t*′ = 0) = **1** can be relaxed. Adjusting *N*_*A*_(*t*′ = 0) allows 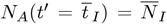 to be preserved. Then both 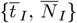 can be treated as model inputs. The 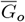 prefactor in Eq. [4.1a] can be modified to give:

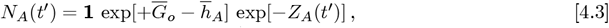

so that 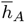 can adjust *N*_*A*_(*t*′ = 0), while keeping the same *t*′ = 0 point:

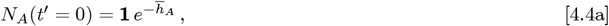

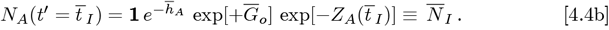

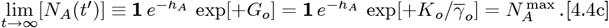

Here 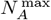 is the total number of CoVID-19 *Resurgence* cases at the pandemic end for this *EIM* model. Given values for 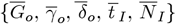, Eq. [4.4b] uniquely sets 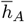 via:

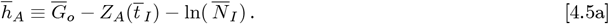

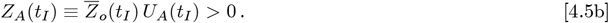

The *N*_*A*_(*t*′) of Eq. [4.3] then gives this *ρ*_*A*_(*t*′):

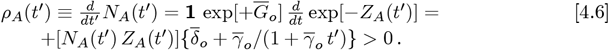

for the daily number of new CoVID-19 cases, providing a self-consistent analytic function for *ρ*_*A*_(*t*′), instead using *ρ* _*A*_(*t*′) ≈ Δ *N*_*A*_(*t*′) / ≈ Δ*t*′ as a numerical approximation. For long times, Eq. [4.6] becomes:

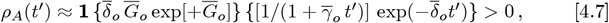

which exhibits a nearly exponentially decaying tail. Minimizing the *rms* error using a *Logarithmic Y-axis* vs linear-time axis gives **Figs. 5-6**, with these best fit parameter values and results:

**Figure 5.**
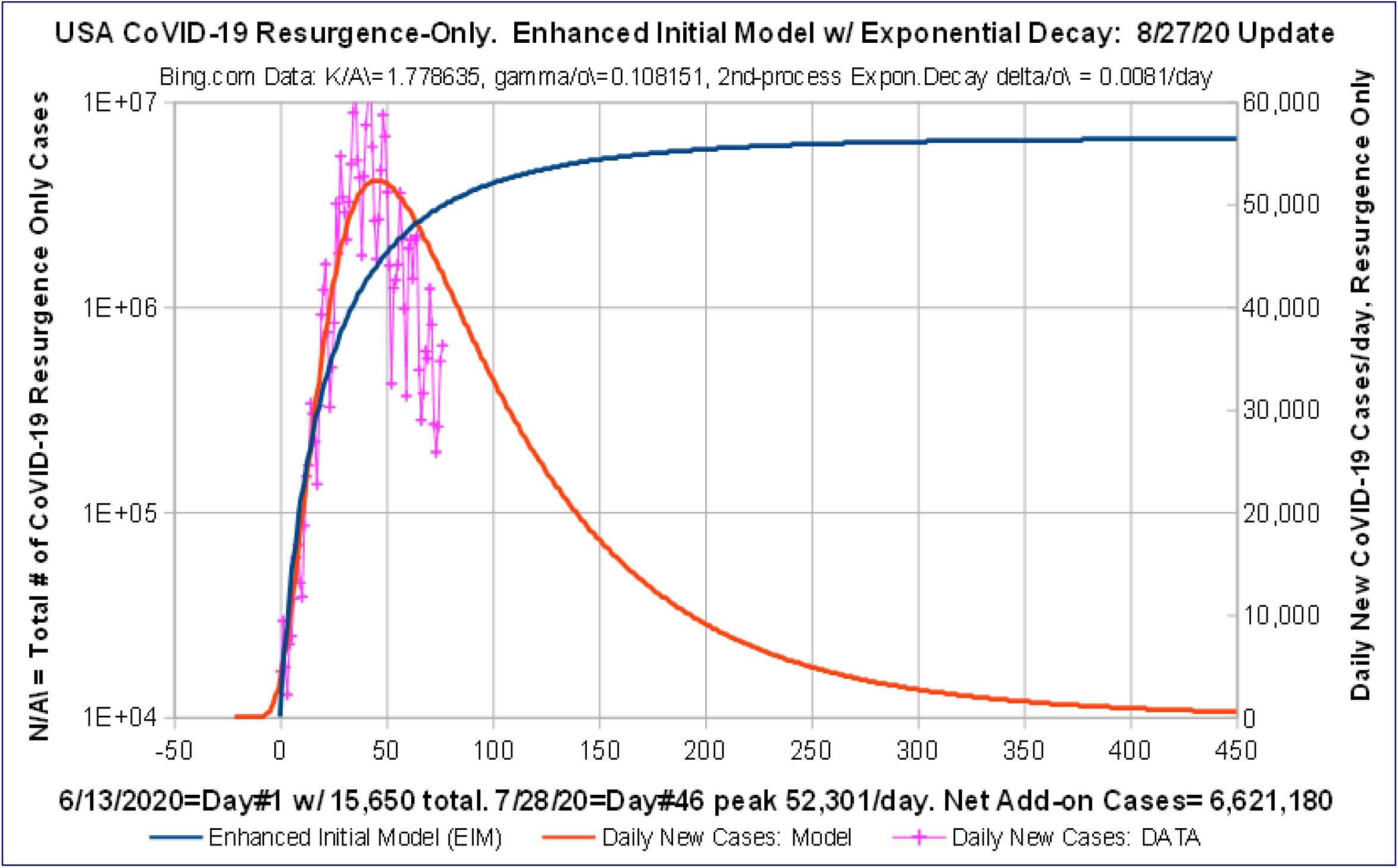
*Enhanced Initial Model (EIM):* USA CoVID-19 *Resurgence* 6/7/20-8/27/20. *EIM* best fit with ***N*_*A*_*[t] ∼ exp(-ZA[t])*** using enhanced ***Z*_*A*_*[t]*** function with exponential decay term, which significantly improves fit. Best fit uses *Logarithmic Y-axis*: *ln(****N*_*A*_*[t]***) vs time.

**Figure 6.**
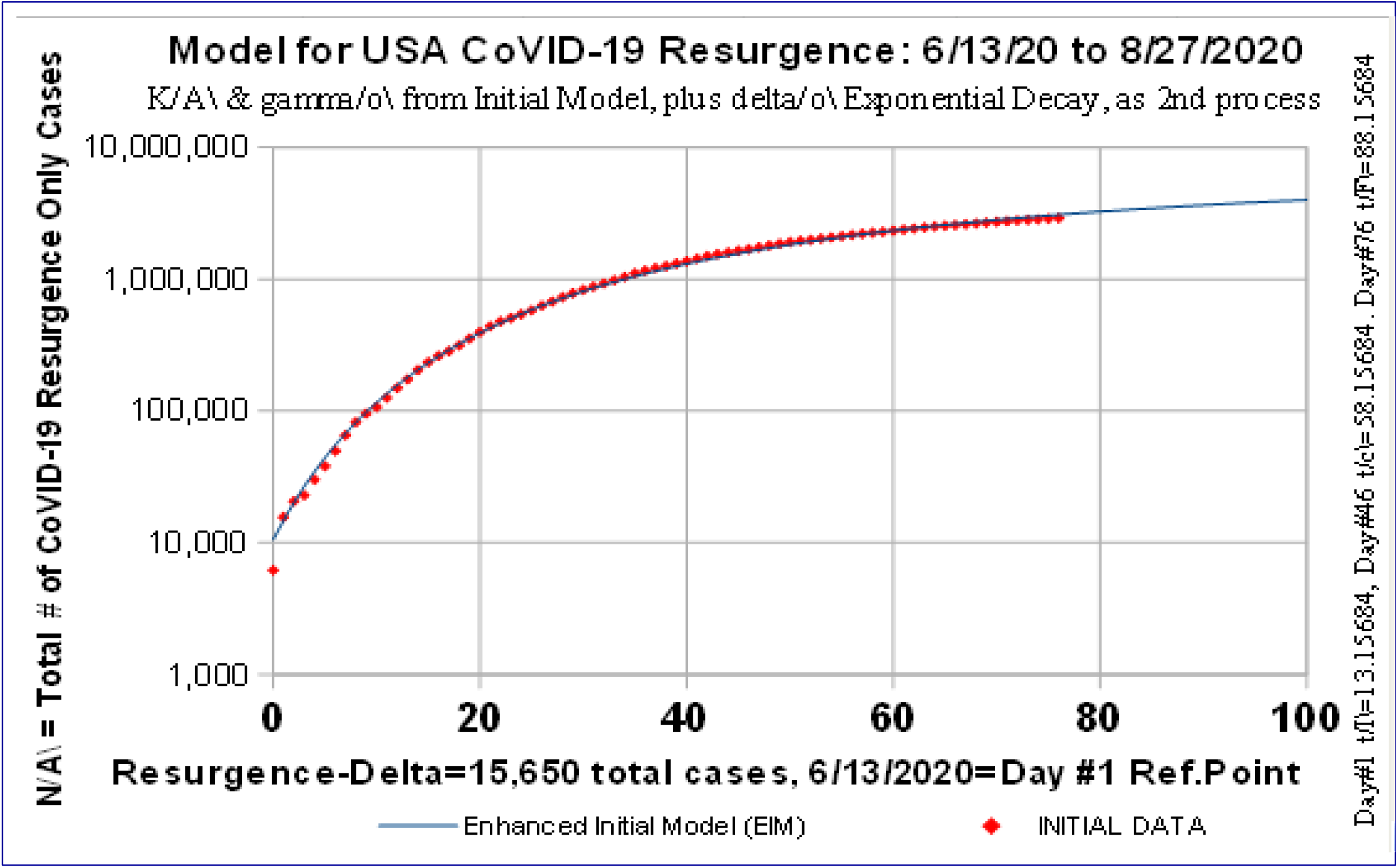
Enhanced *Initial Model (EIM)*: USA CoVID-19 *Resurgence* 6/7/20-8/27/20. *EIM* best fit with *N*_*A*_[*t*] ∼ *exp*(-*Z*_*A*_[*t*]) using enhanced *Z*_*A*_[*t*] function with exponential decay term. Datafit minimizes rms error on *Logarithmic Y-axis*.

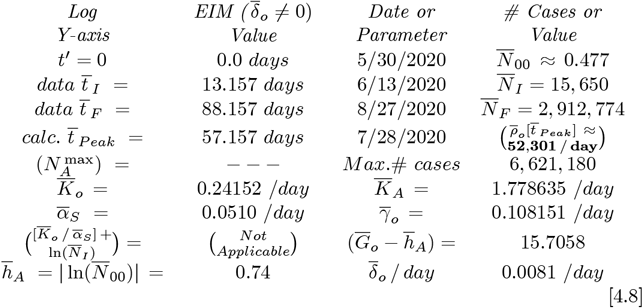

The **Fig. 6** *Logarithmic Y-axis* data fit is quite good, as is **Fig. 5** when compared to the *IRM* **Fig. 3**. The faster decaying **Fig. 5** *ρ*_*A*_(*t*′) tail gives a significantly lower prediction for the total number of *Resurgence cases* at the pandemic end. Finally, a similar data fit is shown in **Figs. 7-8**, except the *rms* error was minimized using a *Linear Y-axis* vs linear-time axis for the *ρ*_*A*_(*t*′) *Resurgence* data. It has the following best fit parameters, which are similar to the above Eq. [4.8] table results:

**Figure 7.**
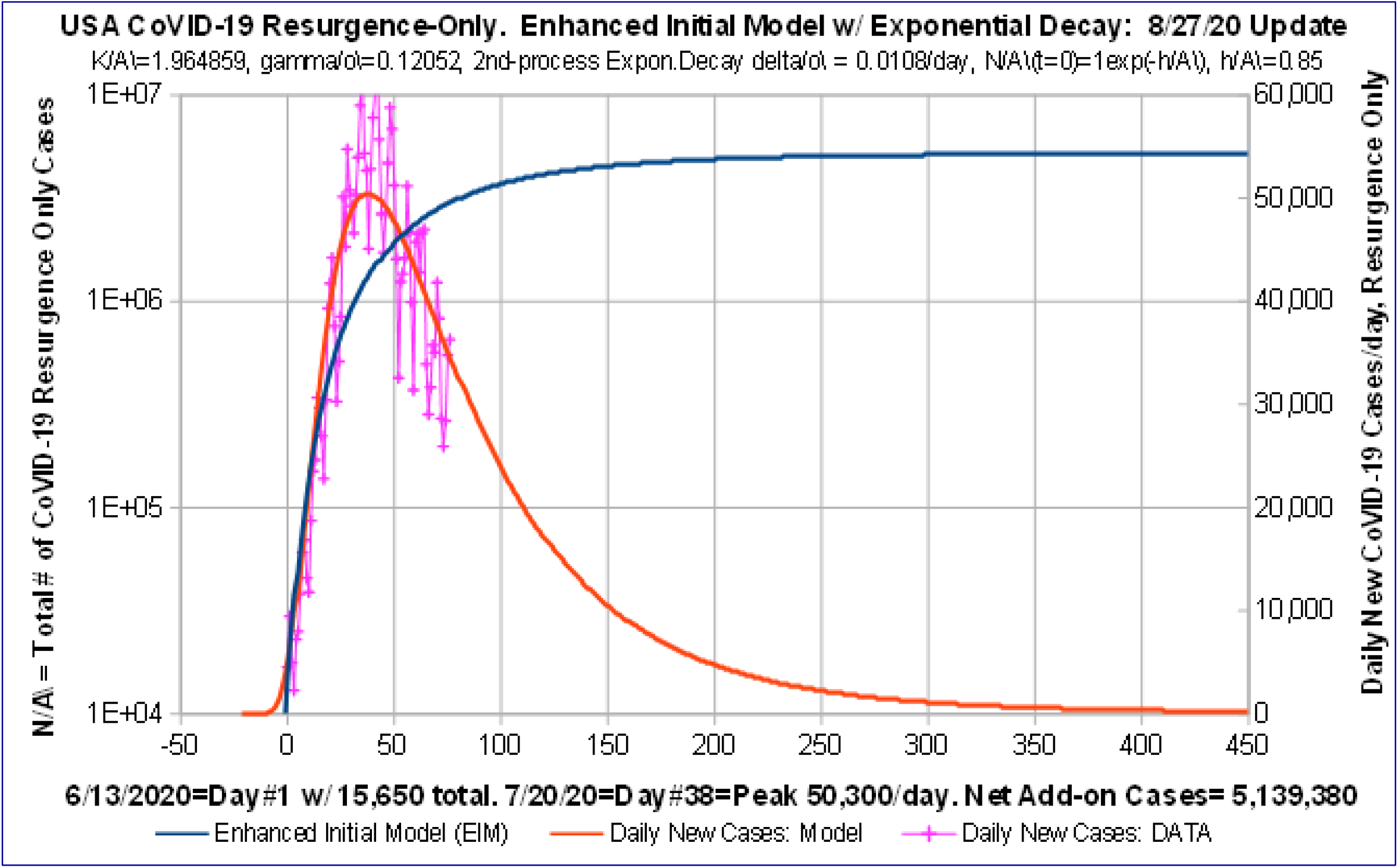
*Enhanced Initial Model (EIM)*: USA CoVID-19 *Resurgence* 6/7/20-8/27/20. *EIM* best fit with *N*_*A*_[*t*] ∼ *exp*(-*Z*_*A*_[*t*]) using enhanced *Z*_*A*_[*t*] function with exponential decay term. Bestfit minimizes error on Linear Y-axis vs time w/ ***Y***= *dN*_*A*_[*t*]/*dt* = Daily # New CoVID-19 cases.

**Figure 8.**
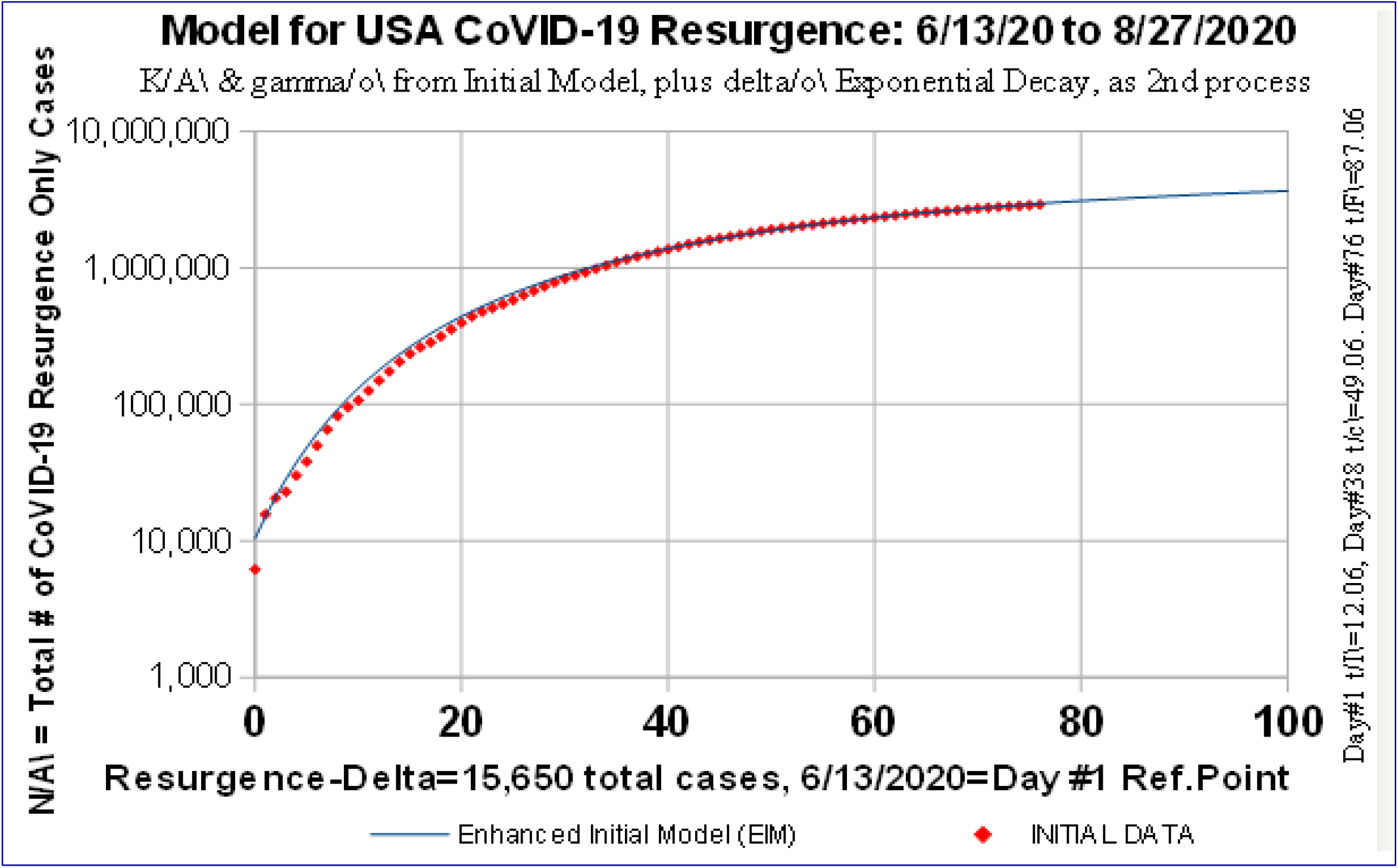
Enhanced Initial Model (EIM): USA CoVID-19 Resurgence 6/7/20-8/27/20. *EIM* best fit with *N*_*A*_[*<u>t</u>*] ∼ *exp*(-*Z*_*A*_[*t*]) using enhanced *Z*_*A*_[*t*] function with exponential decay term. Deviations on *Logarithmic Y-axis* due to minimizing error using *Linear Y-axis* as given in **Fig. 7**.

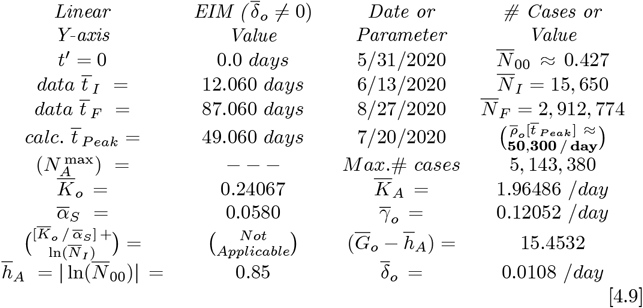

These values form our best estimate for USA CoVID-19 *Resurgence*. Combining these results with the pre-*Resurgence* data fit of **Figs. 1-2**, gives **Figs. 9-10** for the full USA CoVID-19 evolution, covering the entire 3/21/2020-8/27/2020 time frame. This final data fit captures virtually all of the shape nuances in the actual data. The predicted final number of USA CoVID-19 Cases at the pandemic end, from Eq. [2.7] and Eq. [4.9] is:

**Figure 9.**
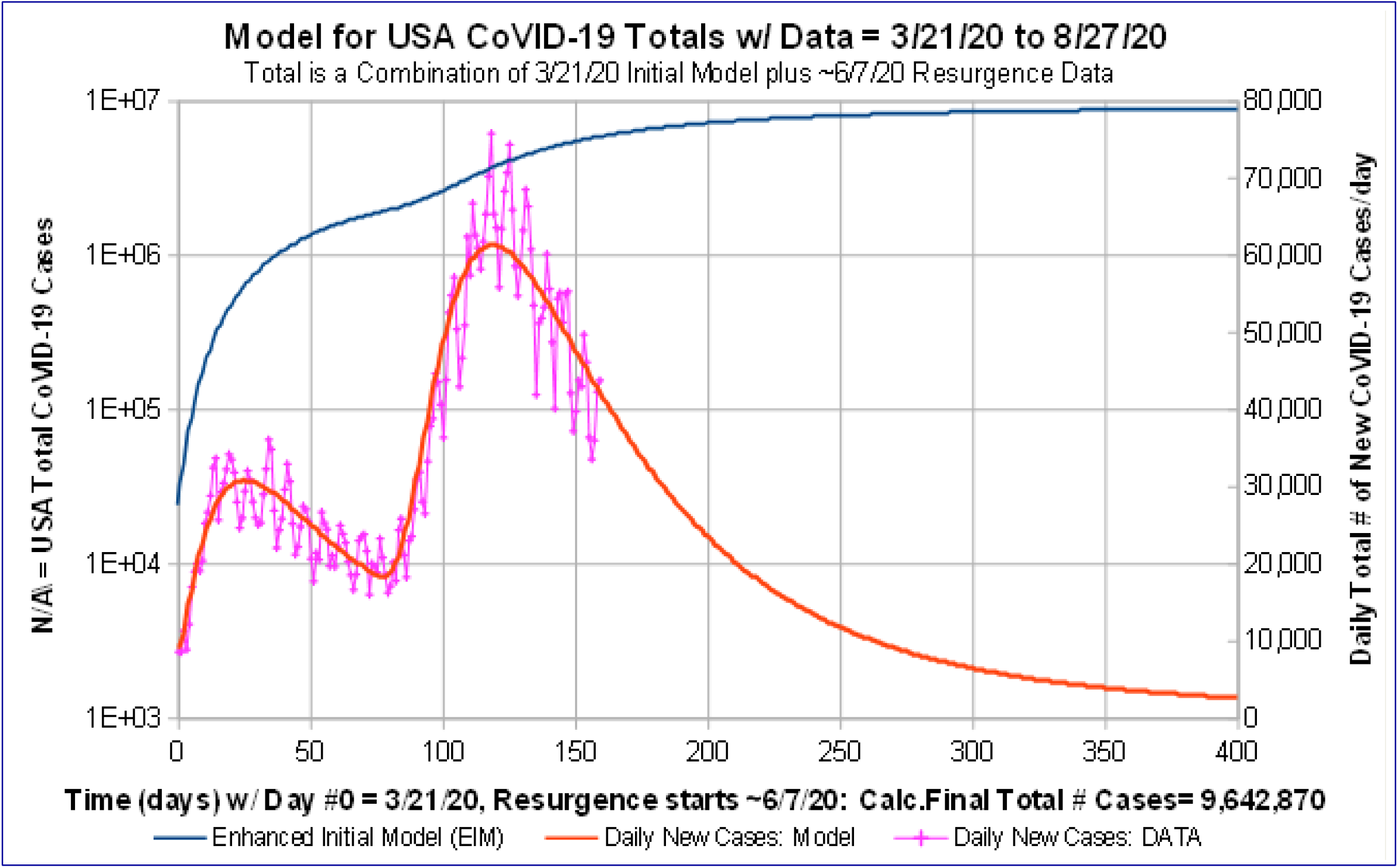
USA CoVID-19 Totals: *IMB* Plus *EIM* for *Resurgence* 3/21/20-8/27/20. Combination of *Initial Model Baseline (IMB)*, starting from 3/21/20 [**Fig. 1**]; plus *Enhanced Initial Model (EIM)* for CoVID-19 Resurgence, starting from ∼6/7/2020 [**Fig.7**].

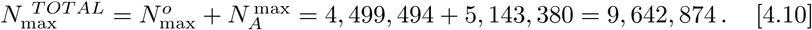

For CoVID-19 in the USA, **Fig. 11** plots the ratio of the total number of deaths versus total number of cases (% vs time), based on the bing.com database^**1**^, which gives ∼2 9325% = (169, 108) / (5, 766, 718), as of 8/27/2020. This value is similar to the IHME 8/27/2020 value^**5**^ of ∼3 1065%, which is shown as a horizontal line on **Fig. 11**.

Using the slightly higher IHME mortality rate allows our **Fig. 10** predictions to be compared with the most recent IHME predictions^**5**^, as shown in **Fig. 12**. The IHME predictions include the presumption of a 2^*nd*^ *Resurgence*, due to factors^**6**^ of “*seasonality and declining vigilance*”. Each IHME projection shown in **Fig. 12** is also an IHME Model average^**5**^, with the magnitude of their lower and upper bound deviations (not graphed) being < 2 5% by 9/26/2020, increasing to < 42% by 1/1/2021. The IHME 2^*nd*^ assumptions are evident in the upward (+) curvature in all IHME predictions, as compared to the downward (-) curvature of the present *Resurgence* model, indicating progress to a CoVID-19 pandemic shut-off, assuming **NO** 2^*nd*^ Resurgence occurs.

**Figure 10.**
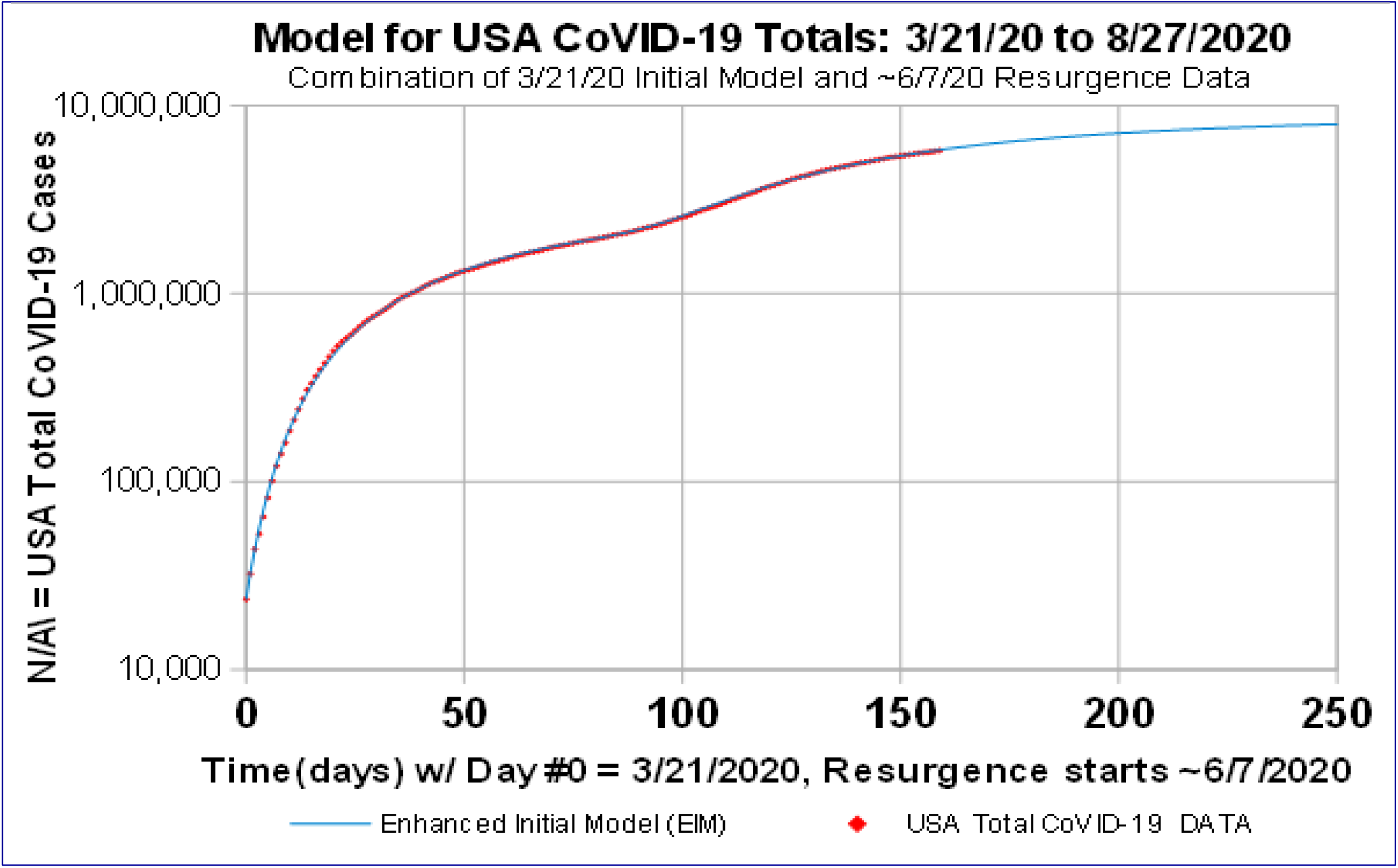
USA CoVID-19 Totals: *IMB* Plus *EIM* for *Resurgence* 3/21/20-8/27/20. Combination of *Initial Model Baseline (IMB)* from 3/21/20 [**Fig. 2**]; plus *Enhanced Initial Model (EIM)* for CoVID-19 *Resurgence* from ∼6/7/2020 [**Fig. 8**] gives **9,642,874** total at pandemic end.

**Figure 11.**
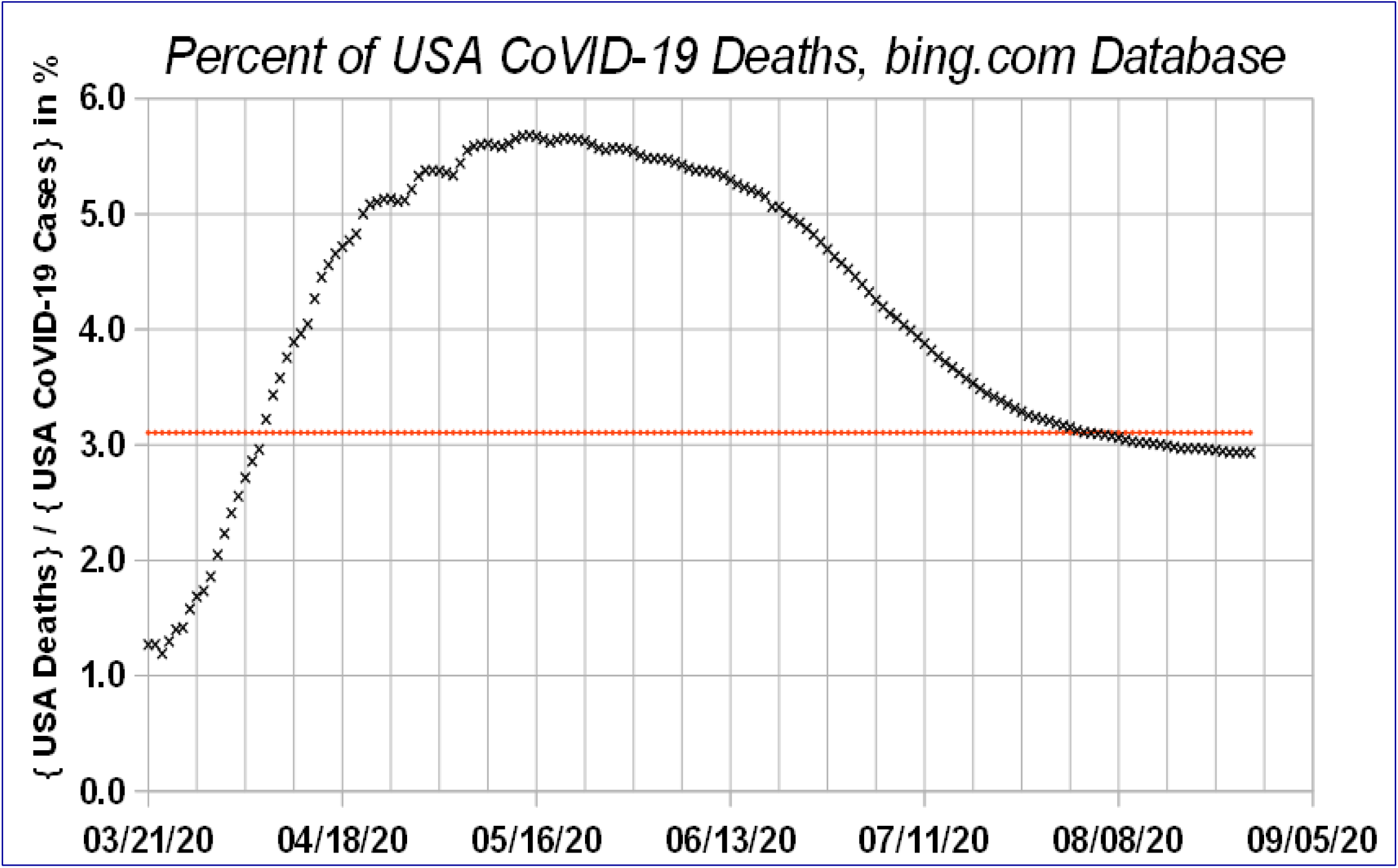
Net Percent *USA CoVID-19* Deaths: Ratio of Total # of USA Deaths to Total # of *bing*.*com* reported USA CoVID-19 Cases, 3/21/20 through 8/27/2020. Some USA CoVID-19 restrictions lifted ∼6/7/2020 leading to CoVID *Resurgence*; IHME used ***3*.*1065%*** as of 8/27/20.

**Figure 12.**
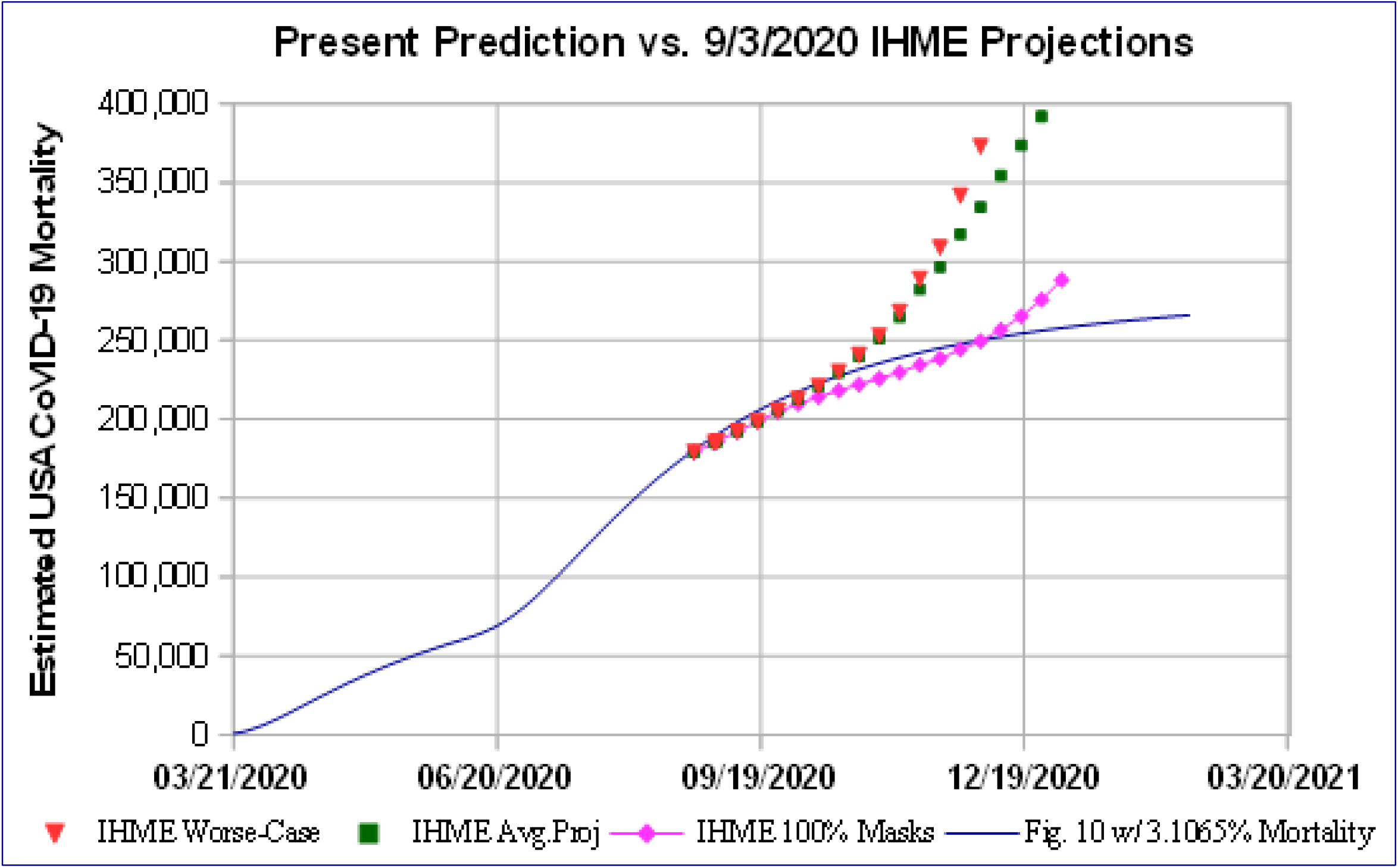
Net Predicted *USA CoVID-19* Deaths: The 8/27/20 IHME ***3*.*1065%*** Mortality Rate for USA CoVID-19 cases was applied to **Fig. 10** to estimate USA CoVID-19 mortality, assuming **NO** *2nd Resurgence;* enabling comparison to IHME’s model, which has *2nd Resurgence* effects.

The causes of a 2^*nd*^ *Resurgence* could include a large-scale set of new re-openings, creating another rapid rise in CoVID-19 cases, similar to **Fig. 9**. A follow-on analysis would be needed for this 2^*nd*^ *Resurgence*. The possibility of multiple CoVID-19 waves was highlighted early on by the University of Minnesota CoVID-19 team^**7-8**^, but each wave was assumed to have **minimal overlap**. Instead, these results, and the IHME projections (which already includes a 2^*nd*^ *Resurgence*), support the idea that USA CoVID-19 evolution is likely to have **multiple overlapping** waves of *Resurgence*.

## 5 Discussion and Conclusions

The *Initial Resurgence Model (IRM)* for USA CoVID-19 *Resurgence*, given by Eqs. [3.3a]-[3.3d] and Eqs. [3.6a]-[3.6b] has the *γ*_*o*_ parameter accounting for the effects of society-wide *Social Distancing*. Our prior work^**2**^ showed that the effects of implementing society-wide shut-downs changed the CoVID-19 pandemic evolution within days of the start of its implementation. Thus, the size of *γ*_*o*_ likely reflects the degree to which society-wide large gatherings were eliminated. It is a non-local parameter that is generally not part of the traditional **SEIR** (*Susceptible, Exposed, Infection, and Recovered or Removed*) pandemic modeling, which are governed by local differential equations.

Our analysis shows that the USA CoVID-19 *Resurgence* data decreased faster than the *IRM* model predictions. A similar situation^**3-4**^ was seen in the CoVID-19 pandemic evolution in Italy, which was successfully modeled by introducing a second process:

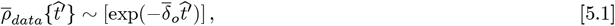

which has an exponentially decaying tail. This second process is independent of the gradually changing *t*_*dbl*_ *doubling time*, which gave rise to the *IRM* 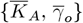 parameters.

For USA CoVID-19 *Resurgence*, an *Enhanced Resurgence Model (ERM)* was developed to include this second process. This *ERM* essentially replaces the 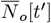 and 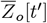 of Eqs. [3.3a]-[3.3b] with:

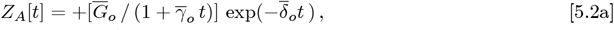

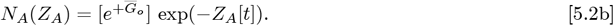

The necessity of using a second process (*δ*_*o*_ ≠ 0) to model the USA CoVID-19 *Resurgence* has a potentially important implication. This *δ*_*o*_ is a second non-local parameter that may not be part of a traditional **SEIR** (*Susceptible, Exposed, Infection, and Recovered or Removed*) pandemic model. Pre- vs Post-*Resurgence*, what else changed? The most likely explanation is that *δ*_*o*_ measures the degree to which people wear and wore masks to mitigate and prevent CoVID-19 spread during the *Resurgence*.

## Data Availability

All data used is in the Public Domain or was on the CoVID-19 website maintained by bing.com.

